# The association of skeletal muscle energetics with recurrent falls in older adults within the Study of Muscle, Mobility and Aging (SOMMA)

**DOI:** 10.1101/2023.11.08.23298267

**Authors:** Philip A. Kramer, Ezequiel Zamora, Haley N. Barnes, Elsa S. Strotmeyer, Nancy W Glynn, Nancy E. Lane, Paul M. Coen, Peggy M. Cawthon, Bret H. Goodpaster, Anne B. Newman, Stephen B. Kritchevsky, Steven R. Cummings

## Abstract

**Background:** Falls in the older population are a major public health concern. While many physiological and environmental factors have been associated with fall risk, muscle mitochondrial energetics has not yet been investigated.

**Methods:** In this analysis, 835 Study of Muscle, Mobility and Aging (SOMMA) participants aged 70-94 were surveyed for recurrent falls (2+) after one year. Skeletal muscle energetics were assessed at baseline in vivo using ^31^P Magnetic Resonance Spectroscopy (MRS) (ATPmax) and ex vivo by High Resolution Respirometry (HRR) of permeabilized muscle fibers from the vastus lateralis (MaxOXPHOS).

**Results:** SOMMA participants who reported recurrent falls (12%) had a slower 400m walk gait speed compared to those with 0-1 falls (1.0 +/-0.2 vs. 1.1 +/-0.2, p<.001) and took a greater number of medication in the 30 days before their baseline visit (5.6 +/-4.4 vs. 4.2 +/-3.4, p<0.05). MaxOXPHOS was significantly lower in those who reported recurrent falls (p=0.008) compared to those with one or fewer falls, but there was no significant difference in ATPmax (p=0.369). Neither muscle energetics measure was significantly associated with total number of falls or injurious falls, but recurrent falls were significantly higher with lower MaxOXPHOS (RR=1.33, 95% CI= 1.02-1.73, p=0.033). However, covariates accounted for the increased risk.

**Conclusions:** Ex vivo maximal muscle mitochondrial energetics were lower in older adults who experienced recurrent falls, but covariates accounted for its association with recurrent fall risk, suggesting this “hallmark of aging” may not be directly implicated in the complex etiology of falls.

## Introduction

Falls in the older population are a leading cause of morbidity and mortality and represent a worldwide public health concern. Falls have consequences beyond physical injury in the older population as they have been associated with reduced quality of life, fear of falling, loss of independence, and institutionalization (1). According to the CDC, 1 out of 4 individuals over 65 years of age experience a fall each year which result in nearly 30,000 deaths (2). Understanding the biological factors that increase risk of falls will likely be helpful in developing effective preventive strategies and interventions.

Falling is complex and results from the interaction of multiple risk factors (3-7). One important fall risk factor is the age-related decline in muscle mass and strength, or sarcopenia, which affects balance, mobility, and physical function (8-13). Mitochondrial dysfunction is a major hallmark of aging, a key contributor to sarcopenia, and has been associated with low leg power and cardiorespiratory fitness, but it is unclear if this association extends to fall risk (14). As the major energy producing organelle in muscle cells, mitochondria are sensitive to metabolic, inflammatory, and oxidative injury associated with obesity, exercise, and inactivity (15). Aging is reported to decrease the mitochondria’s capacity for oxidative phosphorylation (OXPHOS) and ATP production and increase the production of reactive oxygen species (ROS), particularly in high-energy demand tissue like skeletal muscle (16-18). Mitochondria are particularly susceptible to oxidative stress as accumulation of ROS further impairs their function through mitochondrial DNA mutations, affecting mitochondrial respiratory chain, and altering membrane permeability and Ca^2+^ homeostasis (19). It has been proposed that a decline in the OXPHOS capacity of the mitochondria would result in a decrease in muscle power and physical performance, thereby increasing the risk for falls and related injuries.

To examine the role of muscle energetics in fall risk, we analyzed data from the Study of Muscle, Mobility and Aging (SOMMA), using *in vivo* and *in vitro* measures of muscle mitochondrial energetics in 835 older adults over the course of 1 year. Since maximal OXPHOS and ATP generation directly correlate with muscle strength and physical function, (17,20) we hypothesized lower muscle energetics will be associated with increased risk of recurrent falls in this population.

## Methods

### Study Design and Participants

SOMMA is an observational multicenter cohort study that recruited community dwelling adults ≥ 70 years from 2 university medical centers between 2019 and 2022. Approval was obtained by institutional review board WIRB-Copernicus Group (WCG IRB) (20180764) prior to initiation of the study ^21^. Inclusion criteria included participants without dementia or active malignancy, able to walk 400 meters and climb a flight of stairs, no contraindications to muscle biopsy or magnetic resonance imaging, a 4-meter walking speed ≥ 0.6m/s (if unable to complete a 400m walk), and BMI < 40kg/m^2^. Individuals on anticoagulants and antiplatelets (except for Aspirin) were excluded. SOMMA obtained MaxOXPHOS and/or ATPmax measurements on 868 of the 879 enrolled participants. Fall data (baseline history, 6 months, and 12 months) was obtained on 835 participants.

### Data Collection

At baseline, participants were asked to report their recent fall history, or the number of falls they had experienced during the preceding 12 months. Subsequently, fall occurrence was ascertained by questionnaires every 6 months until the end of year one and combined for this analysis. A fall was defined as an unintentional loss of balance resulting in the individual coming into contact with ground or floor or hitting an object like a table or chair. A participant was considered to have recurrent falls if they had 2 or more falls in the year after baseline (compared with 0 or 1 fall in that time period). Participants who fell were asked about the consequences of the fall and if it resulted in injury. The criteria for an injurious fall was a fall resulting in fractured or broken bone, sprain or strain, bruise or bleeding, hit to or injury of head, or some other unspecified injury. Those with injurious falls were asked if medical attention was sought after the fall, and these falls were subcategorized as serious injuries.

Covariates include clinical site (University of Pittsburgh or Wake Forest University School of Medicine), age, BMI (kg/m^2^) derived from measured height (stadiometers) and weight (digital scales), and self-reported race and ethnicity (Non-Hispanic White or Racial or Ethnic Minority), self-reported sex, average drinks per week in past 12m, and total number of medications (0 if none taken in past 30 days). Accelerometry data was collected on all 3 orthogonal axes using an ActiGragh GT9X at 80 Hz worn on the non-dominant wrist for 7 full days. Baseline daily activity was assessed as total activity count per 24-hour day (imputed).

### Muscle Mitochondrial Energetics Assessments

Percutaneous skeletal muscle biopsies of vastus lateralis were performed under local anesthesia. Approximately 10mg of biospecimen was prepared for respiration analysis. Muscle fibers were separated, permeabilized, washed, and placed into Oxygraph 2K (Oroboros Inc., Innsbruck, Austria) respiratory chambers for *ex vivo* assessment of MaxOXPHOS, which represents the maximal oxygen consumption rate in pmol/(s*mg) of permeabilized muscle fibers in the presence of saturating complex I-linked (malate, pyruvate, glutamate) and II substrates (succinate) and ADP. *In vivo* muscle energetics, or ATPmax, which represents the maximal rate of ATP synthesis by oxidative phosphorylation following a ∼30 second bout of isometric leg kicking, was assessed via 31P MRS with a 3 Tesla MR magnet (Siemen’s Medical System— Prisma (University of Pittsburgh) or Skyra (Wake Forest University School of Medicine). Detailed mitochondrial protocols have been previously described (14,22).

### Data Analysis

Covariates considered in the analysis included age, sex, race (non-Hispanic White vs. racial/ethnic minority), baseline history of any fall 12 months prior to enrollment, alcohol status (average number of alcoholic drinks per week in past 12 months), number of prescription drugs (taken in past 30 days), physical activity level (using accelerometry; average total activity count). Differences in participant characteristics stratified by recurrent falls were assessed using t-tests for normally distributed data and a Wilcoxon rank-sum test for skewed data, and categorical variables were assessed by chi-square test, or Fisher’s exact test. Kruskal-Wallis Test was used to test the association between muscle energetics measures and fall frequency and type due to non-normal distributions for MaxOXPHOS and ATPmax. Negative binomial regression models were used to examine the association of 1SD decrease in MaxOXPHOS and ATPmax with total, recurrent, and injurious fall outcomes. Adjusted risk ratios and corresponding 95% confidence intervals were calculated with significance level p<0.05.

## Results

### Participant Characteristics

A total of 879 participants aged 70-94 with were enrolled in SOMMA across two clinical sites, Wake Forest University School of Medicine and the University of Pittsburgh. Of these, 835 (342 were men, and 493 women) had falls data. Covariates considered in this analysis were site, age, BMI, race/ethnicity, drinks per week over the past 12 months, number of medications taken in past 30 days, history of falls in the past 12 months at baseline, physical activity at baseline (**Table 1**). Data were stratified by 1 fall or less (0-1) and recurrent falls (2+). After the first year, 28.7% of participants experienced a fall and 12% 2+ falls. Individuals who experienced recurrent falls had a slower 400m walk gait speed than those who experienced 1 or no falls. They also reported fewer alcoholic drinks consumed per week in the year before baseline, though this was not significant (p=0.054), and significantly more medications taken in the 30 days before baseline. Of the 100 individuals who reported multiple falls in the first year, 63 (63%) also had a history of falls in the year prior to the start of the study. However, only 167 (22.8%) of the 735 individuals who reported 1 or no falls had a history of falls. Injurious falls were reported in over half (55.1%) of all individuals who had recurring falls and in only 47.1% who had only 1 fall. Serious injuries that required medical intervention were reported in 30.9% of all recurrent fallers, but only 17.9% percent of those who reported only 1 fall.

**Table 1.** Participant Characteristics Stratified by Recurrent Falls (0-1 vs. 2+ falls)

| Participant Characteristics | N | Overall<br>(N= 835) | 0 - 1 Falls<br>(N= 735) | 2+ Falls<br>(N= 100) | P value |
| --- | --- | --- | --- | --- | --- |
| Site: Pittsburgh | 835 | 406 (48.6) | 361 (49.1) | 45 (45.0) | 0.44 |
| Age, years | 835 | 76.3 +/- 5.0 | 76.2 +/- 4.9 | 77.3 +/- 5.7 | 0.062 |
| Sex: Women | 835 | 493 (59.0) | 429 (58.4) | 64 (64.0) | 0.283 |
| Weight, kg | 835 | 76.1 +/- 15.1 | 76.3 +/- 15.1 | 74.6 +/- 15.1 | 0.312 |
| Body Mass Index (kg/m<sup>2</sup>) | 835 | 27.5 +/- 4.5 | 27.6 +/- 4.5 | 27.3 +/- 4.9 | 0.582 |
| Racial or Ethnic Minority | 834 | 126 (15.1) | 115 (15.7) | 11 (11.0) | 0.221 |
| Avg Drinks per week in past<br>12m | 822 | 2.8 +/- 4.4 | 2.8 +/- 4.4 | 2.4 +/- 4.3 | 0.054 |
| Total number of medications | 833 | 4.4 +/- 3.5 | 4.2 +/- 3.4 | 5.6 +/- 4.4 | 0.003 |
| History of fall in year before<br>baseline | 832 | 230 (27.6) | 167 (22.8) | 63 (63.0) | <.001 |
| Total activity counts (x100,000,<br>imputed) | 780 | 1.99 +/- 0.6 | 2.00 +/- 0.6 | 1.95 +/- 0.6 | 0.474 |
| Gait speed from 400m walk<br>(m/s) | 835 | 1.1 +/- 0.2 | 1.1 +/- 0.2 | 1.0 +/- 0.2 | <.001 |
| At least one fall in year after<br>baseline | 835 | 240 (28.7) | 140 (19.1) | 100 (100.0) | <.001 |
| Any indoor falls in year after<br>baseline | 835 | 131 (15.7) | 58 (7.9) | 73 (73.0) | <.001 |
| Any outdoor falls in year after<br>baseline | 834 | 160 (19.2) | 82 (11.2) | 78 (78.8) | <.001 |
| Injurious fall in year after<br>baseline | 833 | 120 (14.4) | 66 (9.0) | 54 (55.1) | <.001 |
| Serious injurious fall in year<br>after baseline | 832 | 55 (6.6) | 25 (3.4) | 30 (30.9) | <.001 |

Data shown as n(%), mean +/- SD.

### Lower Ex Vivo Muscle Energetics in Older Adults with More than One Fall

To determine if muscle mitochondrial energetics differed in individuals who had reported recurrent falls (2+) or injurious falls, we compared *ex vivo* and *in vivo* muscle energetics measures, MaxOXPHOS and ATPmax, to participants who reported only 1 or no falls (**Figure 1A**), or no injurious falls (**Figure 1B**). MaxOXPHOS was significantly lower (p=0.008) in individuals who reported more than one fall, but no difference was observed with ATPmax (p= 0.369). There were no differences in muscle energetics in participants who report injurious falls (1+) compared to no injuries.

**Figure 1.**
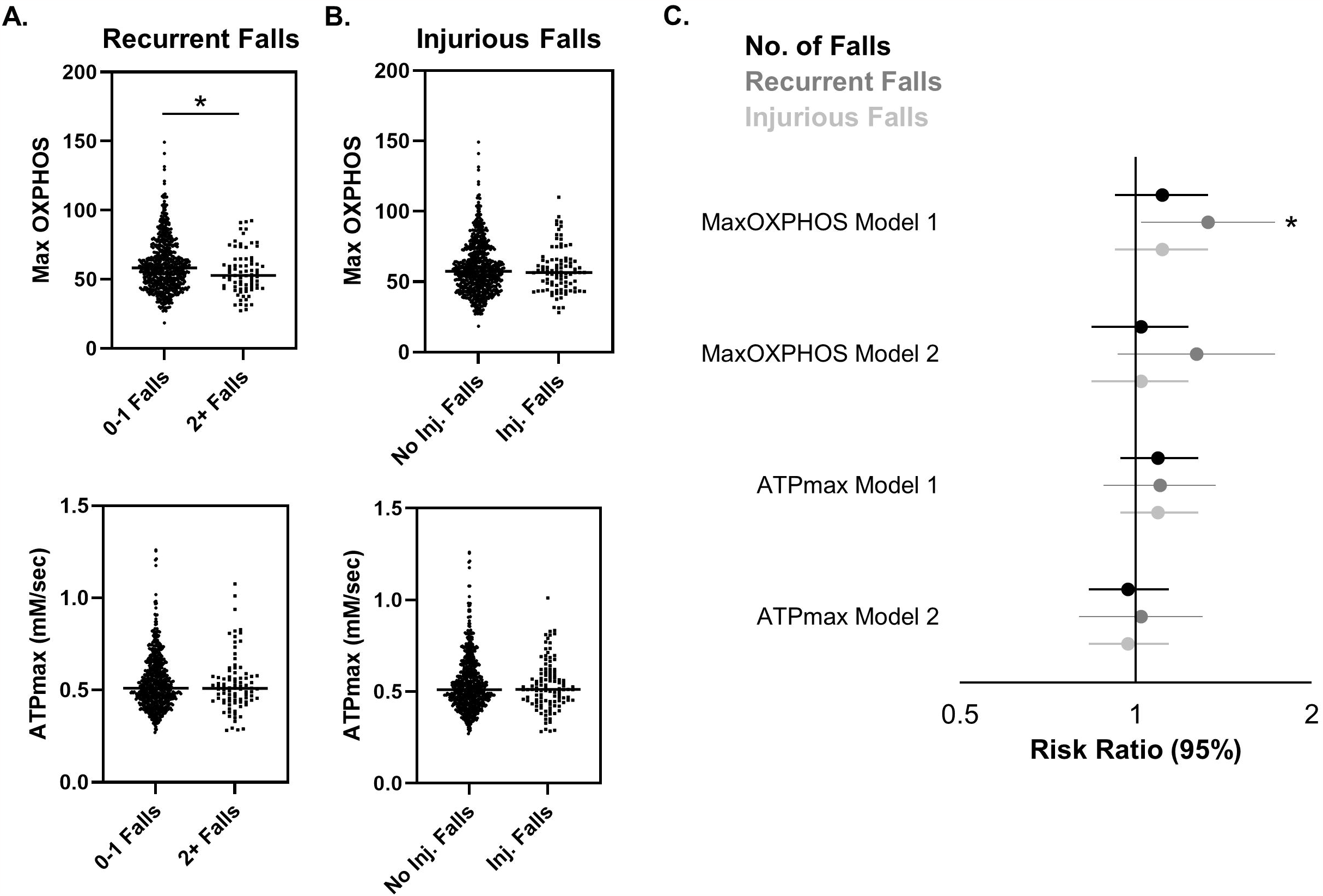
Muscle energetics and risk of recurrent falls. **A)** ATPmax and MaxOXPHOS were assessed in SOMMA participants with 0-1 falls (n=735), and 2+ recurring falls (n=100) and **B)** injurious falls (1+). **C)** A negative binomial regression for total falls, recurrent falls (2+), and injurious falls was performed, and the risk ratio (95% CI) plotted following sequential adjustment for Model 1: Site/technician, Model 2: Model 1 + age, sex, race (white vs. non-white), ETOH status (avg. number of drinks per week in past 12 months), # medications in 30 days before baseline, BMI, total activity counts, and baseline history of falls (y/n). A 1SD change in MaxOXPHOS was equivalent to -18.26 pmol/(s*mg), and ATPmax change of -0.15 mM/s. * = p value <0.05.

### No Significantly Greater Risk of Recurrent Falls with Lower Muscle Energetics

Total number of falls and injurious falls were not associated with changes in muscle energetics (**Figure 1C**), however SOMMA participants had a significant risk of 2 or more falls with lower MaxOXPHOS (RR=1.33, 95% CI= 1.02-1.73, p=0.033), but not ATPmax (RR=1.10, 95% CI= 0.88-1.37). The greater risk of recurrent falls with lower MaxOXPHOS were explained by age, sex, and race, average reported number of alcoholic beverages consumed per week over the past 12 months, number of medications, BMI, physical activity, and baseline history of falls.

## Discussion

In this analysis, 28.7% of participants reported at least one fall and 12% reported 2+ falls during their first year in SOMMA. Those older adults who reported multiple recurrent falls (2+) had significantly lower ex vivo maximal mitochondrial respiration, but not in vivo ATP generation, compared to those who fell once or not at all. The significantly greater risk of recurrent falls due to low muscle energetics was explained by covariates. Recurrent fallers had a modest and borderline-significant (p=0.054) fewer number of alcoholic drinks per week than SOMMA participants who reported 1 or no falls over the course of 1 year. This is interesting as alcohol is known to impair balance in the elderly and increase the risk of falls, however, this might only be applicable in individuals who consume 14 or more alcoholic beverages a week (23). Conversely, recurrent fallers took a greater number of medications in the 30 days before their baseline visit. There is a general understanding that certain prescription medications or a high number of medications, polypharmacy, are independent predictors of fall risk (24). Sex differences in fall risk have also been reported, with 5.6% higher fall risk in women (29.1%) compared to men (23.5%) reported in The English Longitudinal Study of Ageing (25). However, we observed no significant difference in the number of men and women among SOMMA participants who experienced recurrent falls compared to those who did not, though the occurrence of falls in women was 5.1% higher than in men (data not shown). History of falls is a major predictor of future falls (26). This disparity was clearly observed in SOMMA, with 63% of individuals who reported recurrent falls also having a fall history. However, fall history was not associated with lower muscle energetics (data not shown). Future studies into the association of these covariates with muscle energetics are needed.

Falls are a complex, multifactorial, and circumstantial occurrence (25,27,28). Per other reports, fall incidence appear to depend on fall circumstances, including fall location (e.g., indoors, outdoors), type of activity performed in that location (e.g., recreation, housework), and walking surface (e.g., dry, icy) (28,29). In this analysis 73% of recurrent fallers experienced an indoor fall and 78.8% an outdoor fall, while only 41.4% of individuals who reported only 1 fall fell indoors compared to 58.6% who fell outdoors. In SOMMA, we found that outdoor falls were reported with a 2.4% higher incidence than indoor falls in women and a 5% higher incidence in men, with women more likely to report an indoor fall (p=0.04) (data not shown). Women are also significantly more likely to suffer from fall-related injuries compared to men, though obesity, lower muscle strength, and osteoporosis may play a significant role (30). More women self-reported having osteoporosis (27.6%) than men (2.6%); and 85.7% of reported fractures (n=21) at baseline were in women. Due to unreliable self-reporting and possible undiagnosed osteoporosis, we did not account for this covariate in our analysis of injurious falls. However, this suggests injurious falls may not be associated with muscle energetics due to other physiological factors. In this cross-sectional analysis, we were unable to evaluate any survivorship effect, wherein individuals with the poorest of muscle energetics did not survive to later ages or otherwise become ineligible for this study. These exclusion criteria include dementia, malignancy, an inability to walk 400m, walking speed slower than 0.6m/s, or a BMI>40, each of which is reported to associate with muscle energetics (31,32).

Consistent with our hypothesis, recurrent fallers had a lower muscle mitochondrial oxygen consumption (MaxOXPHOS) compared to SOMMA participants with 1 or no falls, and a higher risk of falls was significantly associated with lower muscle MaxOXPHOS. However, this was largely explained by covariates, and ATPmax showed no association. These data suggest that falls are multifactorial and that strategies known to improve muscle energetics could aid in fall prevention, though perhaps indirectly through increased physical activity, lowering BMI, etc. Other interventions may include reducing the use of unnecessary medications or medications known to affect balance. In conclusion, falls alone may not be a suitable outcome in trials that aim to improve mitochondrial function. Future studies are needed to explore other mediators of recurrent and injurious falls to help predict fall risk and develop interventions for older adults.

## Data Availability

All data produced in the present study are available upon reasonable request to the authors.

https://www.sommastudy.com/

## Acknowledgments

The Study of Muscle, Mobility and Aging is supported by funding from the National Institute on Aging, **grant number AG059416**. Study infrastructure support was funded in part by NIA Claude D. Pepper Older American Independence Centers at University of Pittsburgh (**P30AG024827**) and Wake Forest University (**P30AG021332**) and the Clinical and Translational Science Institutes, funded by the National Center for Advancing Translational Science, at Wake Forest University (**UL1 0TR001420**). Author Contributions: P Kramer and E Zamora led the writing team and data meetings. H Barnes, S Cummings, and P Cawthon supported formal analyses. P Cawthon, N Glynn, N Lane, A Newman, E Strotmeyer, and S Kritchevsky provided the most critical reviews and edits that led to the improvement of the manuscript. P Kramer and E Zamora participated in either biopsy collection, processing, and/or experiments. S Cummings, P Coen, B Goodpaster, P Cawthon, A Newman, and S Kritchevsky enabled the study with funding acquisition, project administration, and/or conceptualization of the study.

## Conflicts of Interest

S Cummings and P Cawthon are consultants to Bioage Labs. All other authors report no conflict of interest.

## Notes

### Author Declarations

Approval was obtained by institutional review board WIRB-Copernicus Group (WCG IRB) (20180764) prior to initiation of the study.

